# Automatic Production of Synthetic Labeled OCT Images Using Active Shape Model

**DOI:** 10.1101/2020.09.05.20181917

**Authors:** Hajar Danesh, Keivan Maghooli, Rahele Kafieh, Alireza Dehghani

## Abstract

The challenge of limited labeled data in the field of medical imaging and the need for large number of labeled data for training machine learning algorithms, and to measure the performance of image processing algorithms increases the demand to use synthetic images. The purpose of this paper is to construct synthetic and labeled Optical Coherence Tomography (OCT) data to solve the problems like having access to the accurate labeled data and evaluating the processing algorithms. In this study, a modified active shape model is used which considers the anatomical features of available images such as number and thickness of the layers and their associated brightness, the retinal blood vessels, and shadow information with wise consideration of speckle noise. The algorithm is also able to provide different datasets with varying noise level. The validity of our method for synthesis of retinal images is measured by two methods (qualitative assessment and quantitative analysis).

## 1. Introduction

Optical Coherence Tomography (OCT) is vastly used in the different fields like ophthalmology to provide cross sectional images from the eye. Due to its high resolution, this technique has capability to depict the microstructures of a tissue and to discriminate different layers of biologic tissue. The purpose of retinal image analysis is development of computational and mathematical techniques to help ophthalmologists to diagnose diseases such as diabetes, glaucoma, etc., which may cause changes in thickness layers of the retina and the blood vessels [1, 2]. Different image processing methods have already been designed for analyzing these images, with purpose of segmentation of the anatomical layers, segmentation of blood vessels and noise reduction methods [3-6].To evaluate the performance, limitations and clinical application of mentioned algorithms, the validation phase is mandatory [7]. A common method for validating medical image algorithms is the use of Ground Truth (GT) provided by medical experts. Obtaining GT images annotated by experts is a costly and difficult task. Recently, deep learning and convolutional neural network algorithms are shown to be potent to perform accurately in OCT image analysis, such as classification and segmentation [8, 9]. These techniques require a large amount of annotated data in training phase, access to which is crucial for making correct decisions. Such big annotated dataset is rarely available in the field of medical image analysis. Therefore, synthetic data and augmentation methods have been developed to enhance the variety of medical data [10]. The primary methods for producing medical images were digital phantoms that followed simple mathematical models of human anatomy [11]. Several studies exist for producing synthetic retinal fundus images [10, 12, 13], CT scans [14, 15] and OCT images [16-19] and several studies used synthetic data to compare and measure the performance of image processing algorithms [20-22]. The available literature on production of synthetic OCT images is categorized in Table 1 However, the realistic generation of high-quality medical images remains as an unresolved complexity to current computer vision methods. To the best of our knowledge, there are no public available databases of synthetic retinal OCT images, and providing labeled images for large image dataset is often out of access.

**Table 1.** A review of related work

| Investigator | Data set | OCT system | method | Number of segmentation layer | Segmentation method | Number of synthetic data | Feature added to synthetic image |  |  |  |
| --- | --- | --- | --- | --- | --- | --- | --- | --- | --- | --- |
|  |  |  |  |  |  |  | Blood vessel | noise | pathology and textural information | Intensity of layers |
| Serranho. 2011[16]. | 10 healthy volunteers | Carl Zeiss Meditec, Dublin, CA, USA | Mathematical models | 3(ILM, OSL, RPE) | manual | 10 | - | * | - | * |
| Kulkarni. 2011[19]. | 14 animals | within three different C-Scan volumes | Statistical Model | 7 (NFL-GCL, IPL, INL, OPL, ONL, PRL, RPE) | manual | 14 | * | * | - | * |
| Montuoro. 2014[18]. | 20000OCT scans of over 1000 patients | Spectralis CT, Heidelberg Engineering and Cirrus HD-OCT, Carl Zeiss Meditec | Appearance Models | 2(ILM, RPE) | manual | Not Determined | * | * | - | - |
| Shahrian. 2016[17]. | 5pathologies such as cysts |  | Mathematical Models | 2(ILM, RPE) | manual | 5 | - | * | * | * |

On the other hand, some synthetic images are not constructed based on real data and such fictional images do not describe the features of actual medical images. Making labeled data with high similarity to the reference image is therefore less explored. Furthermore, taking into account the anatomical features of the reference images, such as the number and thickness of the layers and their associated brightness, as well as blood vessels and the noise of the images, creates more realistic data. In our proposed method an automatic segmentation method is applied on OCT images and the results are then corrected by an expert. Such segmented data is fed to our synthetic data production method to make both image and the label simultaneously. In this research, we are looking for a new method to make OCT synthetic images that can overcome the shortcomings in the previous methods and provide an optimal solution set. An improved active shape model is used to construct synthetic data. The further fusion of the information in synthetic images produced by this model enables us to simulate the features of the reference image and apply the various effects of imaging conditions. These synthetic retinal images can be useful for confirming image analysis techniques, like segmentation, denoising, and a wide range of other applications.

This paper is organized as follows. In Section 2, we describe the proposed method for generating the synthetic images. In Section 3 we report the result and our experiments to evaluate them. Finally, in Section 4 we discuss on the proposed method and give concluding remarks and hints for future work.

## 2. Material and Methods

The data used in this paper is obtained from Heidelberg 3D OCT-HRA2-KT machine at Sadra Center of Ophthalmology in Isfahan. The size of each data is N x 512 x 496 voxels, in which N is the number of cross-sectional images of each data, varying according to the quality of the imaging and decision of the ophthalmologist, between the 19 to 65. To make the model, 10 normal volume data (100 B scan images) is used in training satge. The proposed method can be summarized in 6 steps.

### 2.1 Images Preparation For training

The training retinal images are segmented into nine boundaries [23] followed by accurate correction with an expert (Figure!).

**FIGURE 1.**
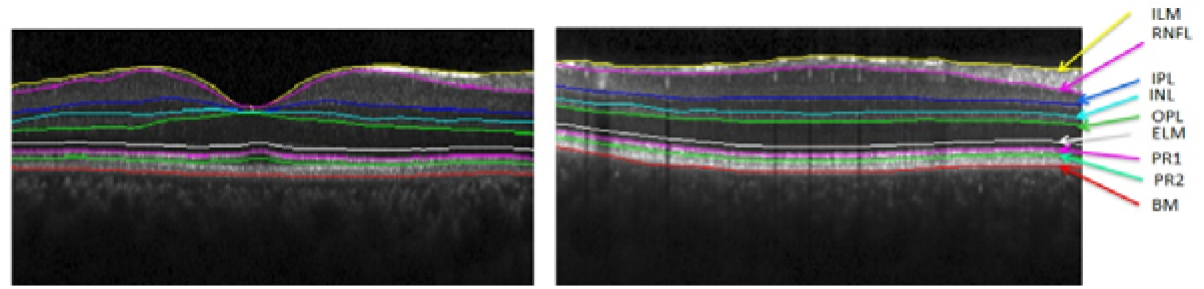
Two samples of training images with automatic segmentation of layers.

### 2.2. Active shape model

The segmented retinal boundaries are then used as training dataset for active shape model [24] to produce new synthetic boundaries. We only incorporate the ASM for data synthesis and don’t use the localization step in this model.

A point distribution model (PDM) is used to describe the shape variation in a training dataset. The shape model is used to construct new shapes similar to the ones in the training dataset. The location of the main points of each retinal layer of the image is determined by analyzing the geometric position of the similar points and construction of a point distribution model. This model includes a mean shape of the location of the points marked in the training stage and a number of parameters for controlling the main modalities of the changes in them. Changes in coordinates of these points describe the change in shape of training dataset. Suppose *x* is the vector of the *n* points of shape *i*

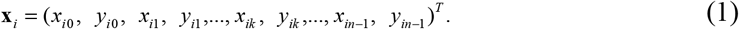

Procrustes analysis [25] is used to align shapes to maximize model specificity and reduce nonlinearity in shape distribution Figure 2 shows the first and last layers before and after aligning.

**FIGURE 2.**
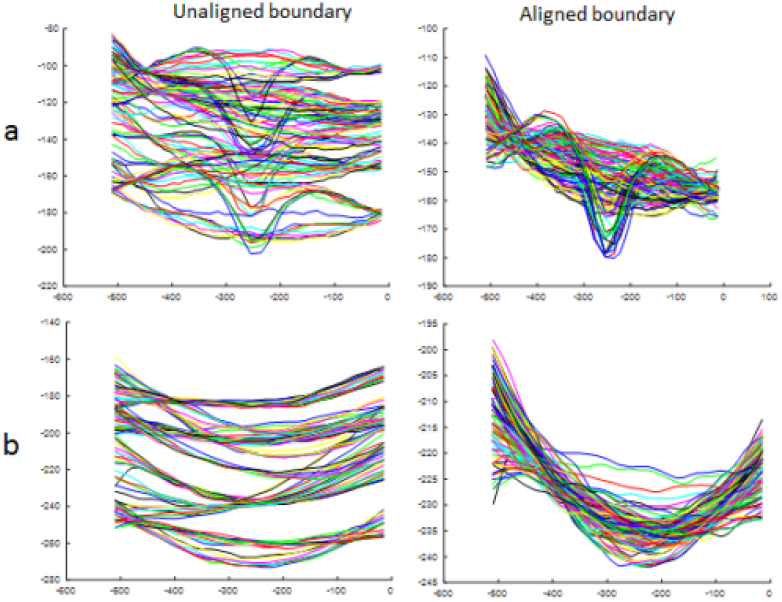
Effects of alignment on boundaries, a) first boundary, b) last boundary.

Principal Component Analysis (PCA) then is applied on the aligned shape vectors. The mean shape of aligned images and the covariance matrix are calculated by [26]:

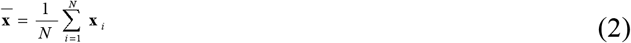

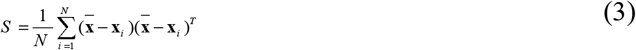

After analyzing the initial component, eigenvectors of covariance matrix are calculated so that the statistical properties of the shape can be obtained. Each shape can be estimated using the following equation:

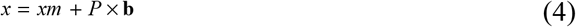

Where *x_m_*, *P* and ***b*** = (*b*_1_, *b*_2_,…,*b_k_*)*^T^*, *k* = (1,…*t*) are the mean shape of geometric location, eigenvectors of the correlation matrix corresponding to largest eigenvalues and a vector of elements (b) containing the model parameters respectively. Equation 4 is the core of our synthetic model and allows us to create a new image by changing the values of *b* vector. These parameters have linear autonomy, although they may be interconnected nonlinearly. The limit on the values of these parameters is also determined by studying the distribution of them in the training images. In the meantime, we can determine a limited range for values to ensure that the parameters change within the permissible range. According to training data, since the changes of *b_k_* in the training set are equal to *λ_k_* (where *λ_k_* is the *k’*th eigenvalue of *S*, the limitation can be defined as follows [27]:

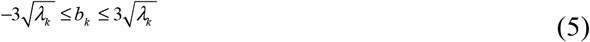

Each aligned sample of the training group shapes is displayed with one point in dimensional space. So, a training group whit *N* image creates a *N* point cloud in 2*n* dimensions space. We assume that this cloud is almost elliptical in shape and its center can be known as the mean shape. The main diameters of the ellipse are obtained by decomposing the PCA on to the images. Each of the diameters represents a mode of change in which the landmarks change around each other to create similar shapes. Eigenvectors of covariance matrices that belong to the largest eigenvalues have produced the largest elliptical shape diameters, thus modeling the most important modes of change in objects. The variations of each eigenvector are equivalent to the eigenvalue corresponding to it, and so only the modes with the highest eigenvalues can be used to describe the changes instead of using all 2*n* available modes.

According to training data and by changing the first four parameters (*t* = 4), similar examples of training shapes can be created. Each of these parameters represents one of the modes of change and models a particular type of variation. However, in our application we forced to put more limitation compared to equation (5) since some parameter values could make visually inconsistent and medically pathologic images. This issue is elaborated in results section.

### 2.3. Finding the most similar reference image to each synthetic image

To transfer some fundamental features such as local brightness, location of the vessels and the choroidal region from reference images to a synthetic image, the most similar image of the training set to the newly created image should be find. For this purpose, after aligning the images the similarity is measured based on the shape of the first boundary. This boundary is therefore divided to 3 sections and the central part is selected as a measure for finding similar B scans. A third-degree curve is fitted to this part and its parameters are extracted, and compared with the available reference images to find the most similar image to the given image.

### 2.4. Applying brightness to synthetic images

After finding the most similar training image to the synthetic image, the average brightness of each layer in selected image is considered as the brightness of the various layers of the synthetic images. In this way, we can create images with different brightness levels which provide more variation in synthetic image and can model the intensity-variation in real images. Figure 3 shows samples from synthetic boundaries and compares the proposed intensity variated method with an easier strategy which obtains the intensities of each layer according to the mean intensity of that layer in training images (global intensity).

**FIGURE 3.**
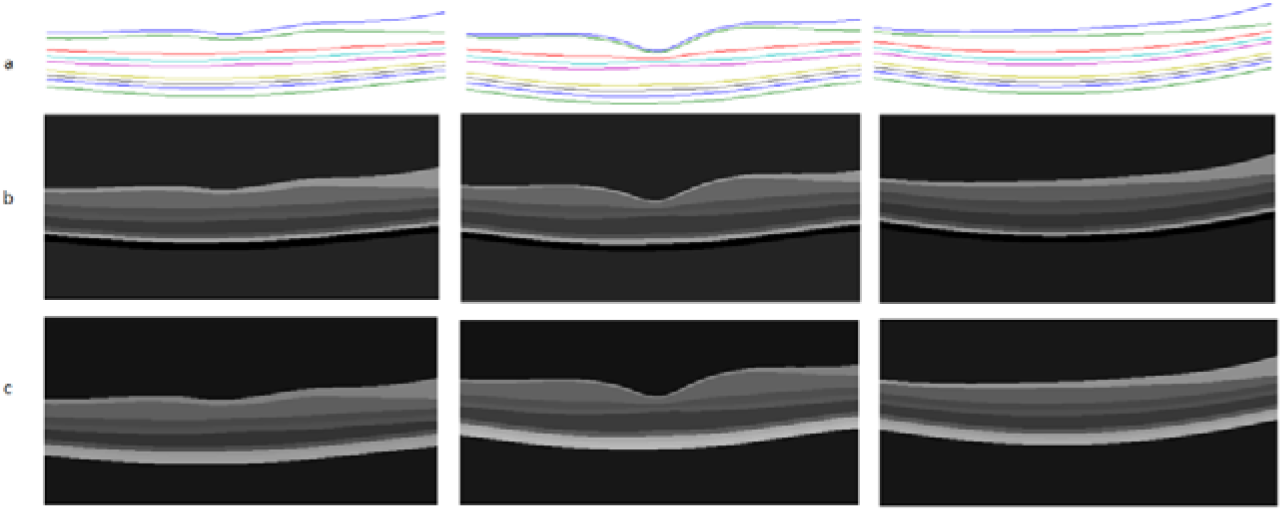
Samples of Applying intensity to synthetic boundaries a) Synthetic boundaries b) Brightness levels selected with global model c) brightness levels selected with intensity variated model.

### 2.5. Adding noise to synthetic images

OCT images are often contaminated with the speckle noise. At this stage, a global speckle noise is applied with various standard deviations and images with different noise levels are created. The noise level can be considerate as a parameter to change the overall quality of synthetic images. Figure 4 shows the synthetic noisy images with different variance levels.

**FIGURE 4.**
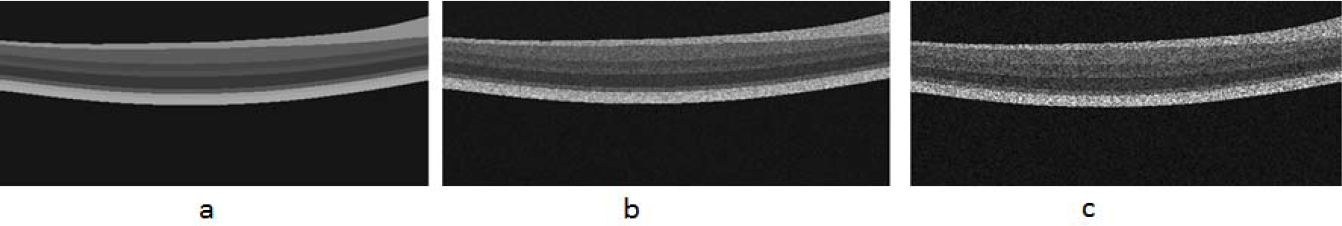
Applying noise to images a) Reference image, b) Adding speckle noise with variance of 0.04 c) Adding speckle noise with variance of 0.2.

### 2.6. Inserting blood vessels

To produce images with highest similarity to the reference image, blood vessels are also added to the image. Since the presence of shadows in the outer layers of the retina is the main factor determining the vessel location [28], after finding the reference image (section 2.3) the brightness profile of the pixels between the boundaries (OPL) and (BM) are computed. A moving average filter is applied to each intensity profile to estimate the drift line to be eliminated. Then a threshold is used to determine the minimum locations, which correspond to location of the vessels. If we select the threshold to be −1.5*std (intensity profile), the location of central point of vessels plus the points around it corresponding to vessel width (v) will be extracted. In next step, we evaluated 3 different methods to add artificial vessels to synthetic image.

In the first method, after locating the vessels, the brightness of the reference image in vessel location is directly copied into the synthetic image. The problem is that in such method the brightness is not adapted to the surrounding brightness values and accordingly, the resulted image may see weird (for instance the inserted vessel might be brighter than the background, which is not common in real images).

To solve this problem, we made a new ASM model for vessel intensity construction (second method). During the training phase mean brightness value of each vessel (with width of V) is selected along with two sets of background brightness (with width of V). Therefore, a brightness profile with length of 3*V* is made for each vessel in training dataset.

By selecting 20 samples of vessels with different widths and resizing them to equal size (9 pixels width), ASM is again used to construct new vessels using the equation 4. For each synthetic image the width and location of the vessels are selected from the reference images, and then synthetic vessels with altered width are placed on these locations.

In third method, we normalized each synthetic vessel (from second method). For this purpose, we considered the local brightness of the location which will host the vessel and normalized the vessel brightness values to match to the host.

Figure 5 shows the location of vessels in a reference image. Figure 6 compares the three methods for inserting blood vessels.

**FIGURE 5.**
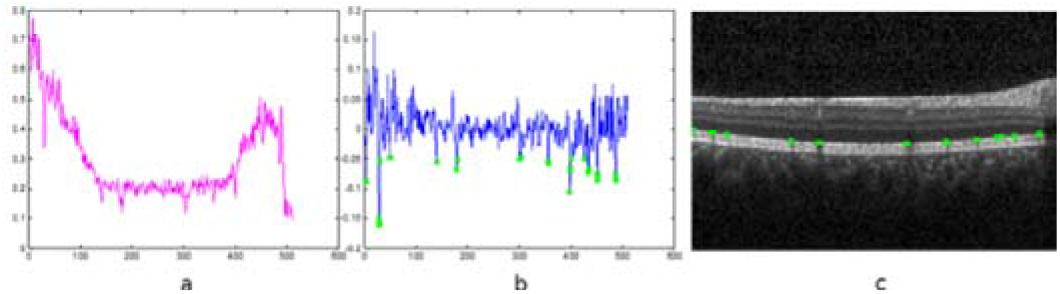
localization process for finding the vessels in a reference image, a) Mean of partial profiles for each column (located between boundaries 7 to 9) b) Applying a moving average filter and threshold c) Location of the blood vessels.

**FIGURE 6.**
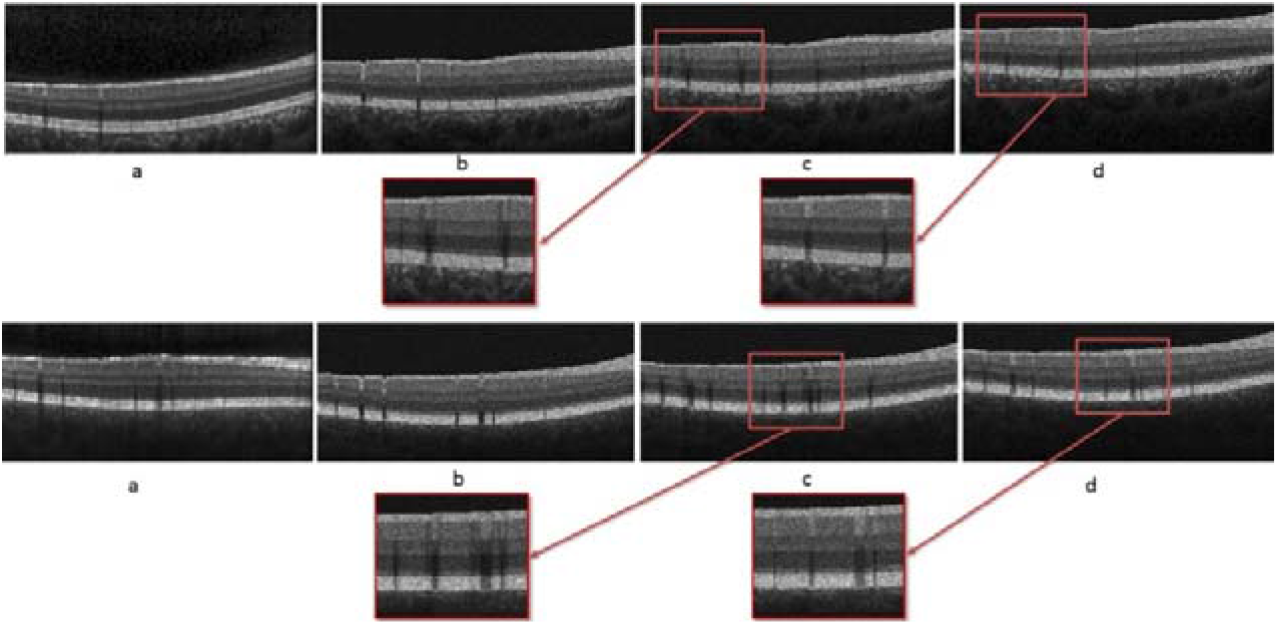
Inserting blood vessels, a) Reference image, b) First method c) Second method d) Third method.

Also designed to make the shape more perfect the choroid region and background of the reference image are transferred to the produced image; this will be visible in the results section.

## 3. Results

In this section we first discuss the limitations that we exposed in selection of *b_k_* for generating normal data. Then we show examples of synthetic data for qualitative evaluation, and finally use segmentation and denoising methodes to evaluate our method quantitatively.

As shown in Figure 7, *b*_1_ parameter is responsible for modeling the overall curvature of RPE and values exceeding 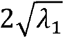 create over shoot on the first and second boundary (weird data); *b_2_* parameter describes the thickness of the layers, (especially RNFL) and weird data is produced for values less than 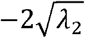*. b_3_* Parameter shows the amount of drooping in macular region and falls out of normal population below the — 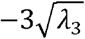 value. *b_4_* Parameter also models the relative thickness of B scans, and its values in the predefined range make acceptable data. As explained in section 2.2 other parameters have more unobtrusive effects, such as angles and curvature shapes. Therefore, we decided that these initial parameters are sufficient for modeling different OCT data.

**FIGURE 7.**
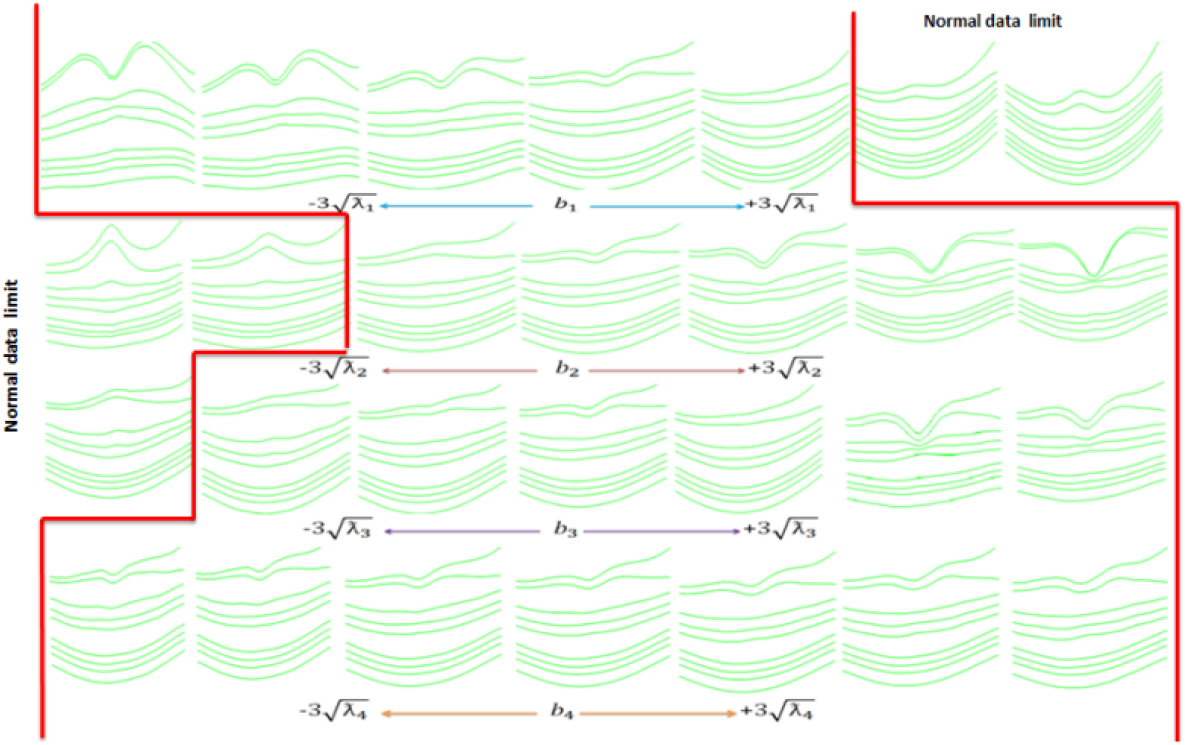
Effects of varying the first four parameters of model.

Sample results of the proposed method for making synthetic OCT images using the features based on the most similar B scans in the training set (reference image) are shown in Figure 8. The reference B scan and the noise free synthetic image are shown in Figure 8.a and 8.b. As shown in Figure 8.c, speckle noise is then added to produce a synthetic noisy B scan. In Figure 8.d, the synthetic model of the blood vessels as well as the choroid layer, are added to the image. The validity of our method for synthesis of retinal images is measured by two methods (qualitative assessment and quantitative analysis).

**FIGURE 8.**
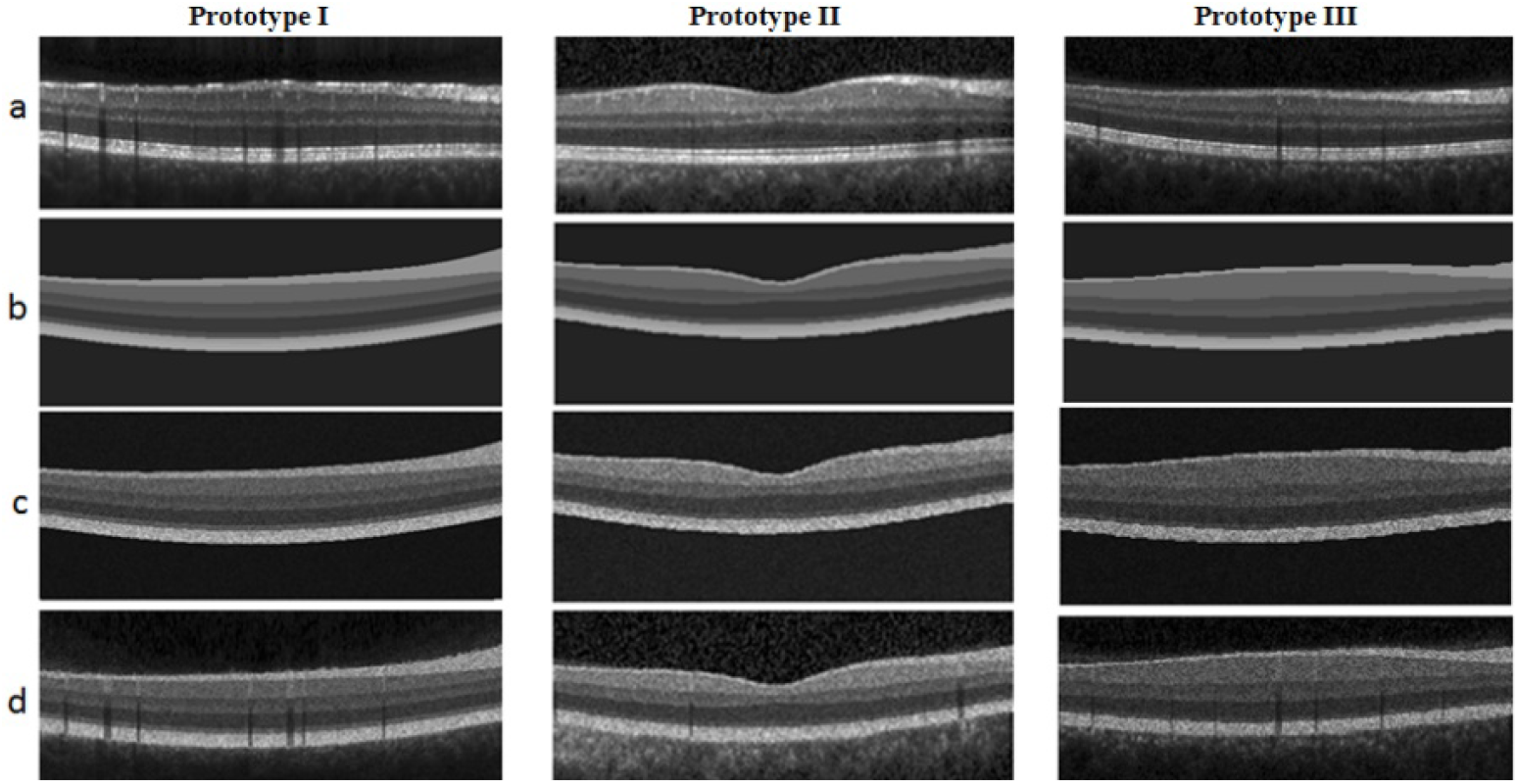
Different stages of Synthetic OCT data for 3 prototypes, a) Reference image, b) Noise free synthetic image, c) Adding noise to the synthetic image, d) Adding synthetic blood vessels.

For qualitative assessment, 40 randomly selected synthetic images were observed by an ophthalmologist to confirm the fallowing properties: the correct order of the blitheness for the layers, the correct shadow effect in lower layers, correct thickening of the RNFL layer in NASAL region, and consistency of choroidal region with properties of the vessels and the layers. All 40 sampled had the mentioned criteria successfully.

To demonstrate applicability of our method in validation of retinal image analysis algorithms, we provide 2 frameworks for quantitative analysis: measurement of segmentation performance and monitoring the denoising performance. In first step, the performance of an automatic segmentation algorithm in different noise levels of synthetic data is tested on 15 randomly selected two-dimensional synthetic data. Figure 9 shows samples of synthetic images with our method and corresponding segmentations. The tested segmentation method is a semiautomatic method based on live wire theory [29].The mean signed and unsigned border positioning errors for each border with different noise levels are presented in Tables 2 and 3.

**Table 2:**
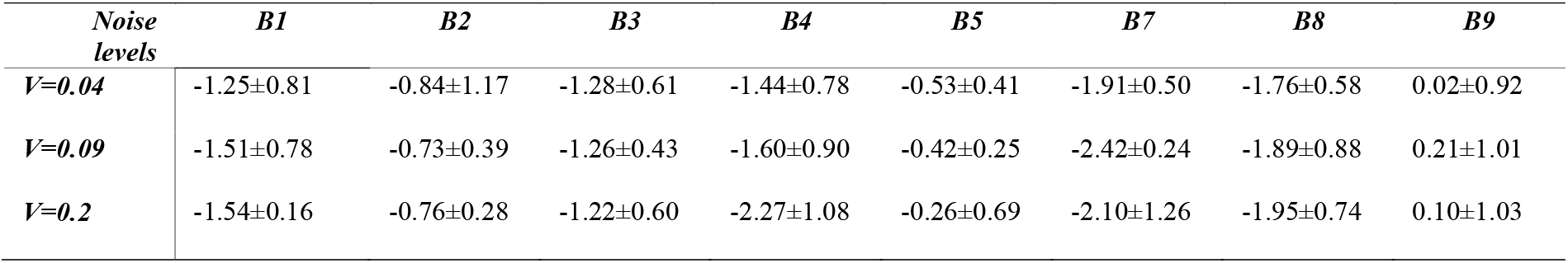
Summary of mean signed border positioning errors (mean ± std).

| Noise levels | *B1* | *B2* | *B3* | *B4* | *B5* | *B7* | *B8* | *B9* |
| --- | --- | --- | --- | --- | --- | --- | --- | --- |
| ***V=0.04*** | -1.25±0.81 | -0.84±1.17 | -1.28±0.61 | -1.44±0.78 | -0.53±0.41 | -1.91±0.50 | -1.76±0.58 | 0.02±0.92 |
| ***V=0.09*** | -1.51±0.78 | -0.73±0.39 | -1.26±0.43 | -1.60±0.90 | -0.42±0.25 | -2.42±0.24 | -1.89±0.88 | 0.21±1.01 |
| ***V=0.2*** | -1.54±0.16 | -0.76±0.28 | -1.22±0.60 | -2.27±1.08 | -0.26±0.69 | -2.10±1.26 | -1.95±0.74 | 0.10±1.03 |

**Table 3:** Summary of mean unsigned border positioning errors (mean ± std).

| Noise levels | *B1* | *B2* | *B3* | *B4* | *B5* | *B7* | *B8* | *B9* |
| --- | --- | --- | --- | --- | --- | --- | --- | --- |
| ***V=0.04*** | 1.89±0.33 | 1.231±0.12 | 1.40±0.51 | 1.55±0.60 | 1.32±0.46 | 1.95±0.39 | 1.81±0.55 | 1.02±0.19 |
| ***V=0.09*** | 1.98±0.27 | 1.26±0.22 | 1.51±0.41 | 2.03±0.68 | 1.10±0.13 | 2.34±0.19 | 2.14±0.63 | 1.25±0.36 |
| ***V=0.2*** | 1.91±0.20 | 1.39±0.15 | 1.56±0.41 | 2.45±1.03 | 1.15±0.25 | 2.15±1.18 | 2.00±0.72 | 1.24±0.36 |

In second framework, we tested a number of famous denoising algorithms to demonstrate the performance of our dataset in evolution of denoising method. BM3d [30] is a baseline algorithm in image denoising which achieved competitive results. We also use the method described in [31] to evaluate the generated images. This method is a numerical optimization framework based on the maximum-posterior estimation of noise-free OCT image and combines a novel speckle noise model, derived from local statistics of empirical spectral domain OCT (SD-OCT) data, with a Huber variant of total variation regularization for edge preservation. This method is expected to exhibit satisfying results in terms of speckle noise reduction as well as edge preservation. The results of using these two methods on 15 randomly selected synthetic images with three different speckle noise levels with variances of (0.04, 0.09, 0.2), are presented with PSNR value in table 4. Furthermore, Figure10 and 11 demonstrates the performance of both methods on two samples of our synthetic data with different noise variations.

**Table4.** PSNR values.

| Speckle noise variance | PSNR dB (BM3d method) | PSNR dB (Li method) | PSNR dB (Nois free synthetic images) |
| --- | --- | --- | --- |
| 0.04 | 36.83 | 37.25 | 27.31 |
| 0.09 | 32.34 | 33.10 | 24.88 |
| 0.2 | 26.11 | 31.56 | 21.48 |

**FIGURE 9.**
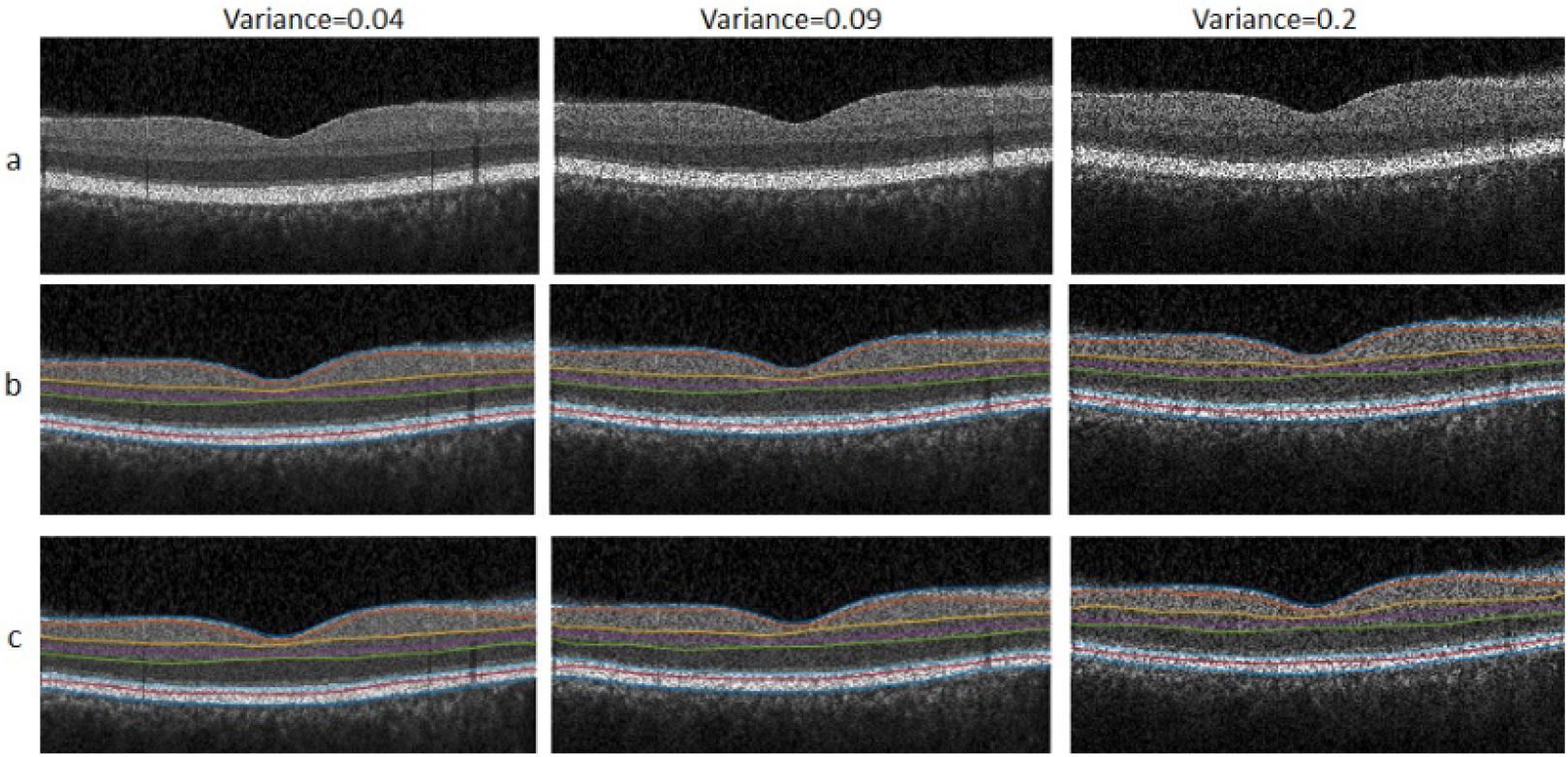
Segmentation results, a) Synthetic images, b) Synthetic boundaries, c) Segmentation boundaries.

**FIGURE 10.**
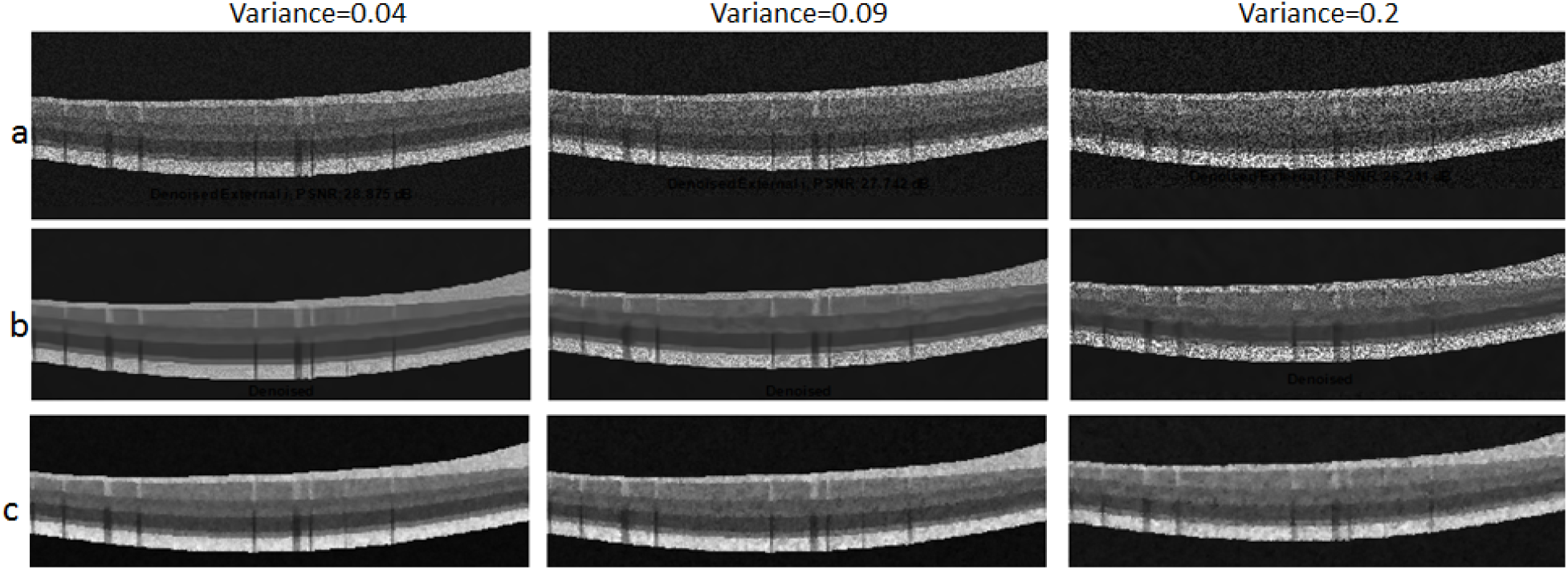
First samples for comparison of denoising methods, a) Synthetic noisy images, b) BM3d method, c) Li method [31].

**FIGURE 11.**
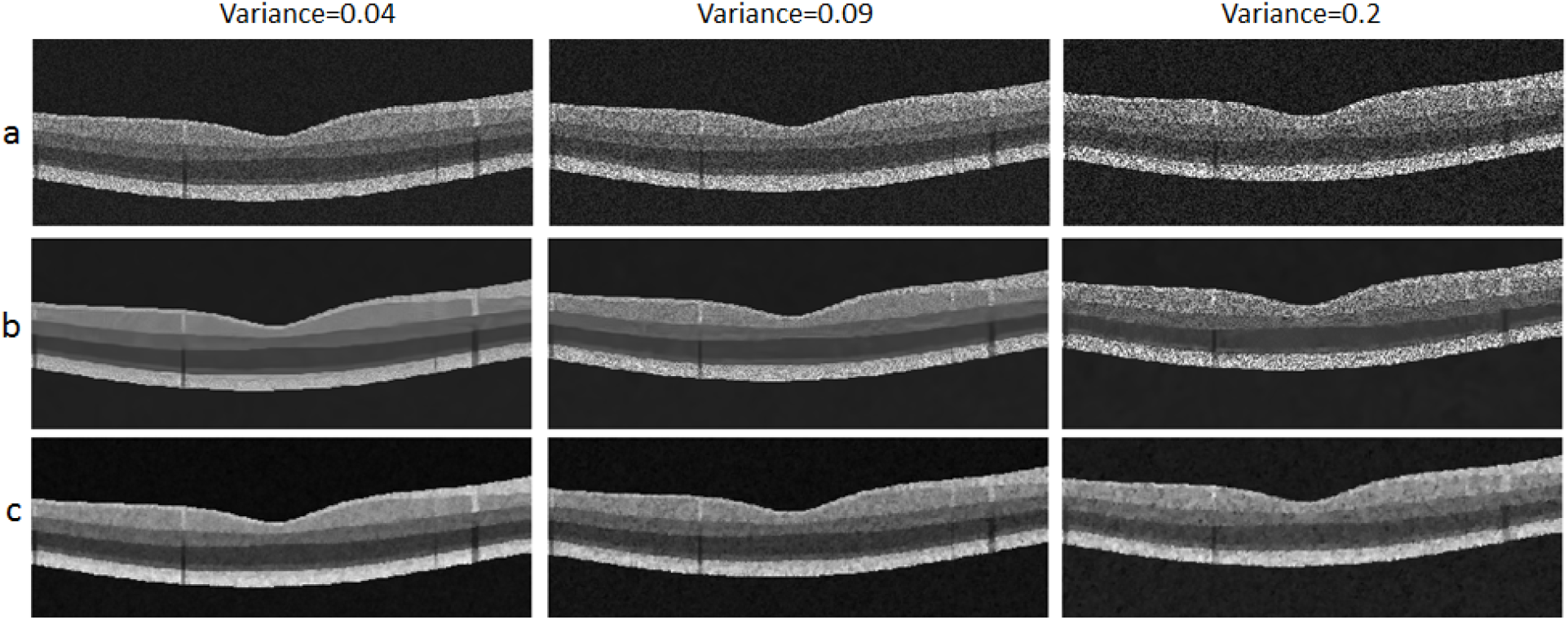
Second samples for comparison of denoising methods, a) Synthetic noisy images, b) BM3d method, c) Li method.

## 4. Conclusion

Unlike other studies on OCT synthesis [16, 17], which only rely on segmented data from internal limiting membrane (ILM), outer segment layer (OSL) and retinal pigment epithelium (RPE), the proposed method is more detailed by incorporating information from nine layers of retina. ASM method is also novel in producing new OCT shape parameters. Furthermore, model construction for making the blood vessels is totally new in this paper. One more emphasized characteristics of the proposed method is production of labeled data which makes it an ideal candidate for performance measurement of segmentation algorithms. Finally, created images can be used to train deep learning algorithms for layer segmentation which need abundant number of labeled data.

## Data Availability

The output data will be available upon to request.

